# Trajectories: a framework for detecting temporal clinical event sequences from health data standardized to the OMOP Common Data Model

**DOI:** 10.1101/2021.11.18.21266518

**Authors:** Kadri Künnapuu, Solomon Ioannou, Kadri Ligi, Raivo Kolde, Sven Laur, Jaak Vilo, Peter R. Rijnbeek, Sulev Reisberg

## Abstract

**Objective:** To develop a framework for identifying prominent clinical event trajectories from OMOP-formatted observational healthcare data.

**Methods:** A four-step framework based on significant temporal event pair detection is described and implemented as an open-source R package. It is used on a population-based Estonian dataset to first replicate a large Danish population-based study and second, to conduct a disease trajectory detection study for Type 2 Diabetes patients in the Estonian and Dutch databases as an example.

**Results:** As a proof of concept, we apply the methods in the Estonian database and provide a detailed breakdown of our findings. All Estonian population-based event pairs are shown. We compare the event pairs identified from Estonia to Danish and Dutch data and discuss the causes of the differences.

**Conclusions:** For the first time, there is a complete software package for detecting disease trajectories in health data.

## INTRODUCTION

Electronic health records are increasingly used for research. They provide a great opportunity for conducting large-scale studies of different diseases and populations that would not be feasible in classical clinical trials or cohort studies. One topic of interest in recent times has been the identification of temporal disease sequences (trajectories) where one event leads to another. This could not only describe progressions within the population but also predict future illnesses from the existing ones.[1-6] An impressive number of temporal relations have been published in trajectory studies, providing a good characterization of these datasets. However, it is difficult to estimate which of these trajectories reflect local healthcare factors such as diagnosis and treatment practices unique to the local or regional healthcare system and which are generalizable globally. In order to find clinically relevant information about disease trajectories that are independent from a particular database and could potentially improve patient care, trajectory studies need to be replicated across a wider database network. The absolute number of disease trajectory studies has remained relatively small. We think this is because of two reasons - first, there is a lack of syntactic and semantic interoperability of health data which makes network studies a challenge, and second, there has not been an open-source standardized implementation of an analytical framework for performing this type of analysis.

The first issue is currently being tackled by various research communities. The open-science Observational Health Data Sciences and Informatics (OHDSI) network has put a tremendous amount of effort into building an open community standard OMOP (Observational Medical Outcomes Partnership) common data model (CDM). OMOP CDM uses standardized vocabulary that transforms data from disparate observational health databases into a common format. This allows the development and wider use of standardized tools for the analysis of electronic medical records regardless of the original formatting of the data.[7] As of today, it is estimated that observational health data of 810 million distinct patients in over 330 databases are partially mapped to the OMOP model,[8] and a wide range of studies have already been conducted on these datasets by a large OHDSI community.[9] Using the same network for investigating temporal health event sequences would enable us to conduct trajectory studies on an unprecedented scale.

Common principles for disease trajectory studies are needed to standardize such studies. We have found that the methods used in previous publications are described insufficiently for adequate replication in other datasets, making it almost impossible to verify the results or conduct a similar analysis in other settings.

In this paper, we propose a standardized framework for detecting prominent temporal clinical event trajectories in the observational health dataset, based on the previous publications and best practices of that field. It is a stepwise process starting with identifying the simplest elements of the trajectories, followed by building longer trajectories of these elements and counting the actual event sequences on that graph. We also introduce the implementation of the framework as open-source software Trajectories that utilizes the OMOP CDM and standardized vocabularies.

## METHODS

### Framework for detecting temporal health event trajectories

The proposed framework for detecting temporal health event trajectories consists of the following steps (**Figure 1**):

**Figure 1.**
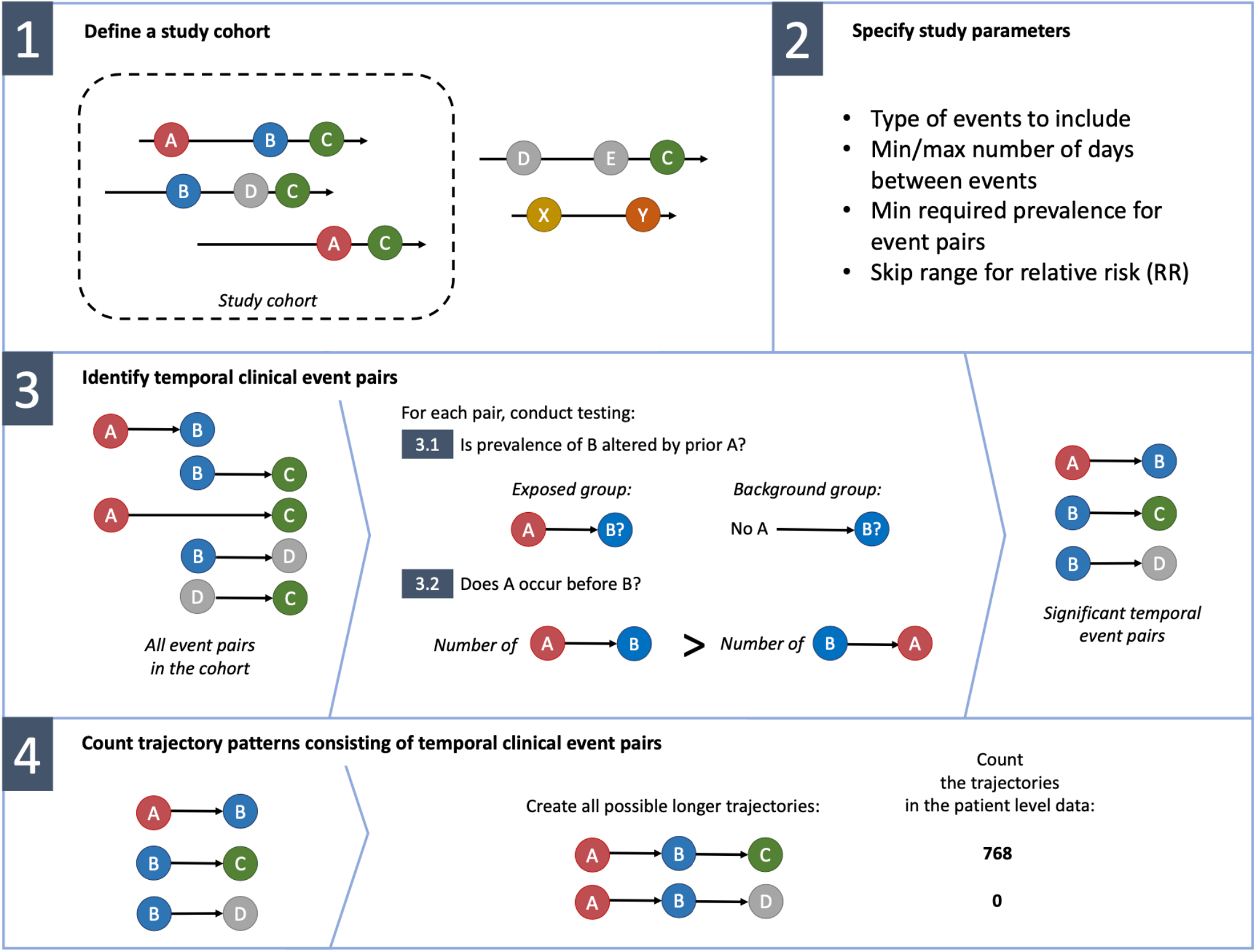
Illustration of the framework.

1. Define a study cohort
2. Specify study parameters
3. Identify temporal clinical event pairs
4. Count trajectories consisting of temporal clinical event pairs

### Define a study cohort

Depending on the research question, disease trajectories can be investigated either in the whole dataset or within a more specific cohort. For example, one may be interested in revealing specific treatment patterns used within a specific cohort such as Type 2 Diabetes, ignoring clinical events related to any other patient group. Therefore, it is vital to clearly define the study cohort at the beginning of any disease trajectory analysis. In our framework, we use flexible and powerful cohort definition principles from OHDSI/OMOP network, where a cohort is a set of persons who satisfy one or more inclusion criteria for a duration of time.[10] These principles have been effectively used in a number of studies across the world,[8,11] allowing for detailed descriptions of the cohorts by using basically any kind of recorded health information. Note that a full database can also form a cohort. Identifying underlying disease pathways from a full database can discover unknown relationships in individuals and timeframes not excluded by the cohort definition.

### Specify study parameters

Within the cohort, there are many additional criteria for the trajectories that need to be specified according to the exact research question.

First, the investigator has to decide which types of clinical events are included in the analysis. While previous trajectory studies have mainly focused on diseases, we extend this approach to any event type that is recorded in observational data. Particularly, a clinical event in the context of our framework is any condition, observation, drug (era), or procedure as determined by the investigator. Within OMOP CDM all the events are coded using standardized OMOP vocabularies.

For discovering ordered temporal sequences where one event leads to another, only the first occurrence of any event for every patient within the cohort is considered, allowing us to avoid counting repeatedly the records with the same (potentially chronic) conditions. This could be a limiting factor for some studies where repetitions of the events play a role - for instance, if one is investigating the sequence of the same type of events or dynamics of numerical measurements associated to same terms. However, taking into account all occurrences of each event when conducting a study is often an impractical approach. For studying diseases that could actually independently occur several times (e.g., seasonal influenza), shorter time frames when defining the cohort should be used allowing the patient to be represented in the cohort with several time periods.

For each patient in the cohort, the selected events form a sequence of events. Any event along that sequence can be considered as a potential risk factor for future events. While some events increase the risk of the future events (relative risk RR>1), others may decrease it (RR<1). For many event pairs, the effect can be very small (RR close to 1) and provide little practical interest. Therefore, depending on the research question, the investigator can specify the range of relative risk of interest, leaving the event pairs that do not satisfy the RR range criteria out of the analysis.

Finally, there are a few parameters that can be used for fine-tuning the analysis. To focus on the most prevalent event sequences, the investigator can set a minimum prevalence for event pairs in order to include them in the analysis. Sometimes it can be useful to limit the minimum and maximum temporal distance in days between the events, preventing events that are either too close or too far apart to be included in the analysis.

### Identify temporal clinical event pairs

Although each individual sequence consists of a number of events, only a few of them have a significant effect on the following events and provide interest for the trajectory studies. The aim of this step is to identify the building blocks for creating longer trajectories. We identify event pairs where the first event tends to not only occur before the other but also alter the risk of the following event. This approach is similar to what has been used by Jensen et al. and Siggaard et al.[1,3]

We perform extensive statistical significance testing of event co-occurrence in event sequences. First, events of each individual sequence are arranged into event pairs - every two events occurring in specific order (may have intermediate events between) within a timeframe specified in the study parameters, form an event pair E1 → E2. Next, for each pair that satisfies all other study parameters described in the previous section, it is assessed whether the first event alters RR of the following event and whether they have a significant temporal order.

For the first task, an exposed (patients having prior E1) and background group (matched patients from the whole cohort) are composed, and the prevalence of E2 in both groups is assessed. For example, Jensen et al. matched exposed and background groups by gender, age group, type of hospital and week of the E1 occurrence in the Danish dataset.[1,3] The number of matching patients will quickly become very small with high levels of stratification, especially for rare events. This would require an initial database of enormous size, as was the case in Denmark.[1] Therefore, our proposed framework requires exact matching by gender, age group and a year of the occurrence of E1 only. Other components are combined into propensity scores and matched. Next, statistical testing is performed. The framework uses Fisher’s exact test to first assess whether the prevalence of E2 in the exposed group is significantly different from the background group.

For all event pairs that demonstrate a significant association, we assess whether there is a significant temporal order (direction) between E1 and E2 in the data using a binomial test (similar to Jensen et al.[1]).

For both tests, the multiple testing corrected p-value below 0.05 is considered statistically significant. We propose to use the false discovery rate (FDR) correction for discovery studies and the more conservative Bonferroni correction for validation studies.

### Count trajectory patterns consisting of temporal clinical event pairs

In the previous step, significant temporal event pairs - the building blocks of longer event trajectories - were identified. These can be further used to form a directed graph where each event is represented as a node and directional edges represent the temporal order between events. The graph is useful for illustrating the main trajectories within the database, especially when less frequent pairs are filtered out (**Figure 2**). The significant temporal event pairs are combined into all possible longer trajectories (e.g., E1 → E2 and E2 → E3, producing the trajectory E1 → E2 → E3) and their actual occurrences are counted by evaluating them against the database. The process is described in detail by Jensen et al.[1] As a result, the list of all trajectories together with their occurrence counts is obtained. The list can be later filtered to answer questions such as how many patients have a trajectory from A to B to C.

**Figure 2.**
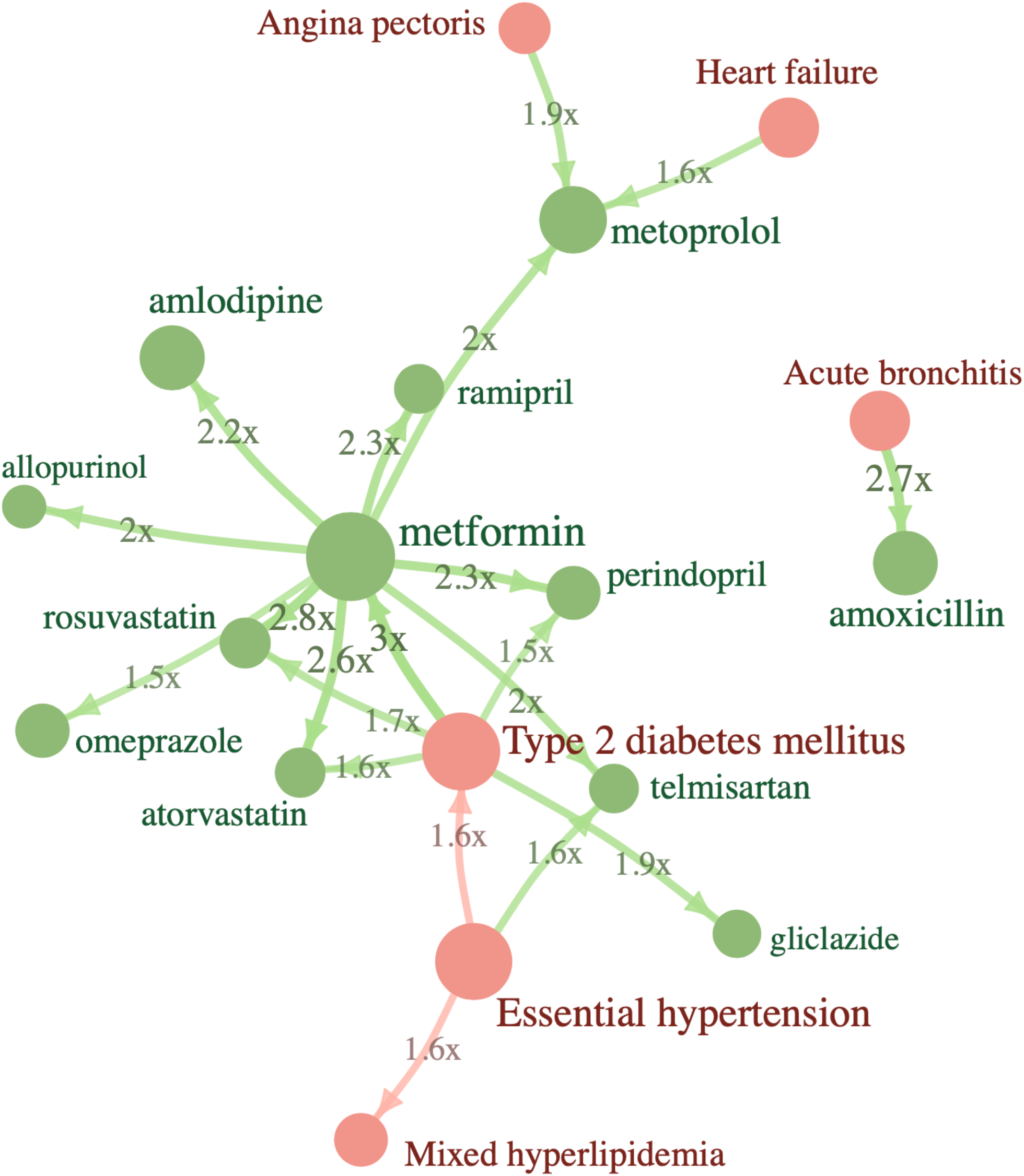
Twenty most frequent event pairs in Type 2 Diabetes cohort in Estonian dataset. Node size indicates the number of patients of that event record, relative risk of the future event is shown on edges. All pairs were also validated as significant in the IPCI database.

### Implementation of the clinical event pair detection framework

The framework described above is implemented as an open-source R package freely available in GitHub^1^, and is open for the community to add further improvements and additional features in the future. It can be run in two modes - either to discover event trajectories from the dataset without any prior knowledge or to validate the event pairs that were discovered in some other dataset. The only difference is that in the validation mode the exact event pairs for testing are given as input. Detailed information on how to run the R package is described in the vignette located in the repository.

Optimal pair matching was performed using the ‘MatchIt’ package, which calls functions from the ‘optmatch’ package.[12-13] For large databases, one can also use faster nearest neighbor matching.

We demonstrate the framework and the package by replicating the largest published trajectory study[3] (Danish population) on Estonian population-based data. In addition, we conducted a Type 2 Diabetes trajectory study on Estonian and Integrated Primary Care Information (IPCI) databases from the Netherlands. The Estonian dataset contains health data of a 10% random sample of the Estonian population (n=147K patients). For each individual in the dataset, all insurance claims, digital prescriptions and in- and outpatient electronic health records from the period 2012-2019 were first converted to OMOP CDM. Mortality rates in the dataset are not complete, covering approximately only two thirds of all deaths. IPCI is a Dutch database containing the complete medical record of more than 2.8 million patients provided by more than 450 general practitioners (GP) geographically spread over the Netherlands. In the Netherlands, all citizens are registered with a GP practice which acts as a gatekeeper in a two-way exchange of information with secondary care. The medical records can therefore be assumed to contain all relevant medical information including medical findings and diagnosis from secondary care. The International Classification of Primary Care (ICPC) is the coding system but diagnoses and complaints can also be entered as free text. Prescription data contain information on product name, quantity prescribed, dosage regimens, strength, indication, and ATC codes.

## RESULTS

### Validation of Danish temporal event pairs in the Estonian population

We analyzed 40,711 temporal event pairs reported by Siggaard et al.[3] as having significant temporal order in the Danish population and tried to confirm or reject these in Estonian data. Both datasets use The International Classification of Diseases version 10 (ICD-10) codes, the Danish dataset being much larger with 7.2 million patients and spanning 24 years (1994-2018). Out of all pairs tested, 13.5% did not occur in Estonian data at all (**Figure 3**). For instance, code “K64” (hemorrhoids and perianal venous thrombosis) is one of the events in 147 temporal pairs in Denmark but the code is never used in Estonia. There physicians still record “I84” instead (Haemorrhoids), although the particular code was removed from ICD-10 in 2010 already.

**Figure 3.**
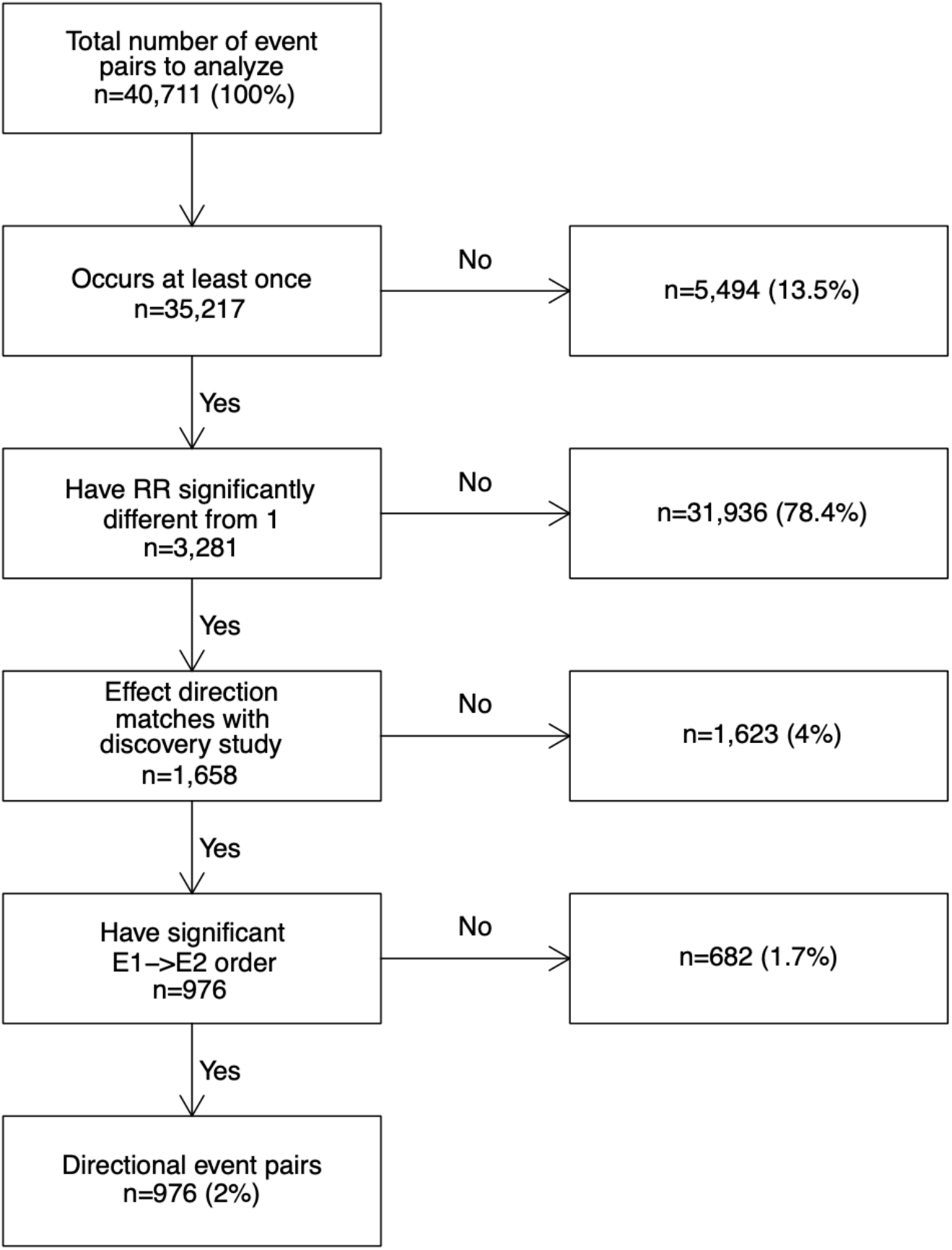
Process flow of testing Danish directional event pairs in Estonian dataset.

For the majority of the pairs tested (78.4%), RR of the future event was not found significantly different from 1 in Estonia. However, such a high number was not a surprise as the Estonian dataset was 49x smaller in patient count and 3x in the time range. The mean counts of these non-significant pairs were 835 in Danish and 46 in the Estonian dataset. Differences in event frequencies play a role here. For example, code “O83” (other assisted single delivery) is frequently used in Danish data (n=59,868 patients) and produced 185 significant temporal event pairs as a result (most prevalent pair “E66” overweight and obesity → “O83” occurred on 10,927 patients). However, in Estonia, the usage of “O83” is extremely rare (n=12) and only 29 of the pairs tested containing that code occurred at least once which is far from being sufficient for observing any statistical significance.

For many pairs where the preceding event altered the RR of the future event, the direction of the risk (increased or decreased) or the order of the two events was opposite of what had been reported in Denmark (4% and 1.7% of the cases respectively). All these pairs had an increased risk in the Estonian data, while a decreased effect was reported in Denmark. An extreme example is “J35” (chronic diseases of tonsils and adenoids) which had a protective effect against future “K83” (other diseases of biliary tract) in Denmark (RR=0.30) but increases the same risk significantly in Estonia (RR=4.5, 95%CI 2.41..8.40).

As a result, we were able to confirm significantly altered RR and temporal order of 976 pairs (2.4%) (**Figure 3**). All validation results are given in **Supplementary File**.

### Discovering event pairs in Estonian data

To discover all temporal event pairs in Estonian data, we ran our package on the whole data without any prior knowledge of Danish findings. We used similar parameters to get comparable results (required pair count >=20). In total, 130,137 event pairs were tested in the Estonian dataset. Out of these, 22,618 pairs in between 797 individual events were found directional and significant (**Figure 4**), but only 4937 pairs of them (22%) overlapped with the Danish directional pairs. What is more, for 2290 pairs (10%), the effect direction of the first event of the pair was similar to the Danish study.

**Figure 4.**
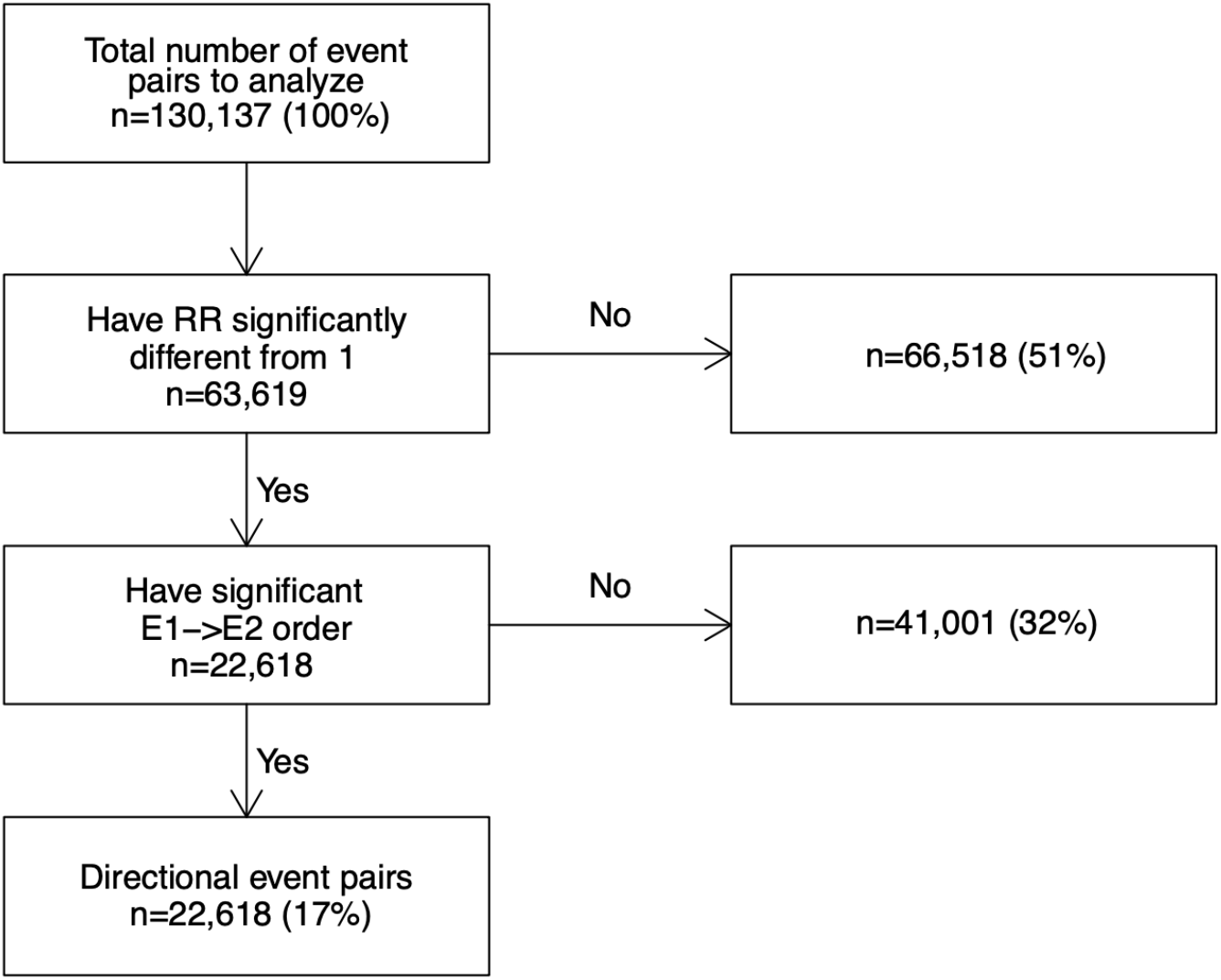
Attrition diagram of identifying directional event pairs in Estonian dataset.

Again, differences in the frequencies of the ICD-10 codes play an important role here (**Figure 5**). For example, code “J06” (acute upper respiratory infections of multiple and unspecified sites) is extensively used in Estonia - recorded for 36% of the patients - leading to 432 temporal event pairs containing “J06” either as the first or the second event. In contrast, only 1.2% of Danish people have a “J06” record and just 15 temporal event pairs are found. Another example is “I11” (hypertensive heart disease) which has a frequency in Estonia 24% vs. 0.5% in Denmark and temporal pair counts 425 vs. 44. Such big discrepancies in individual codes may immediately affect many temporal pairs of events.

**Figure 5.**
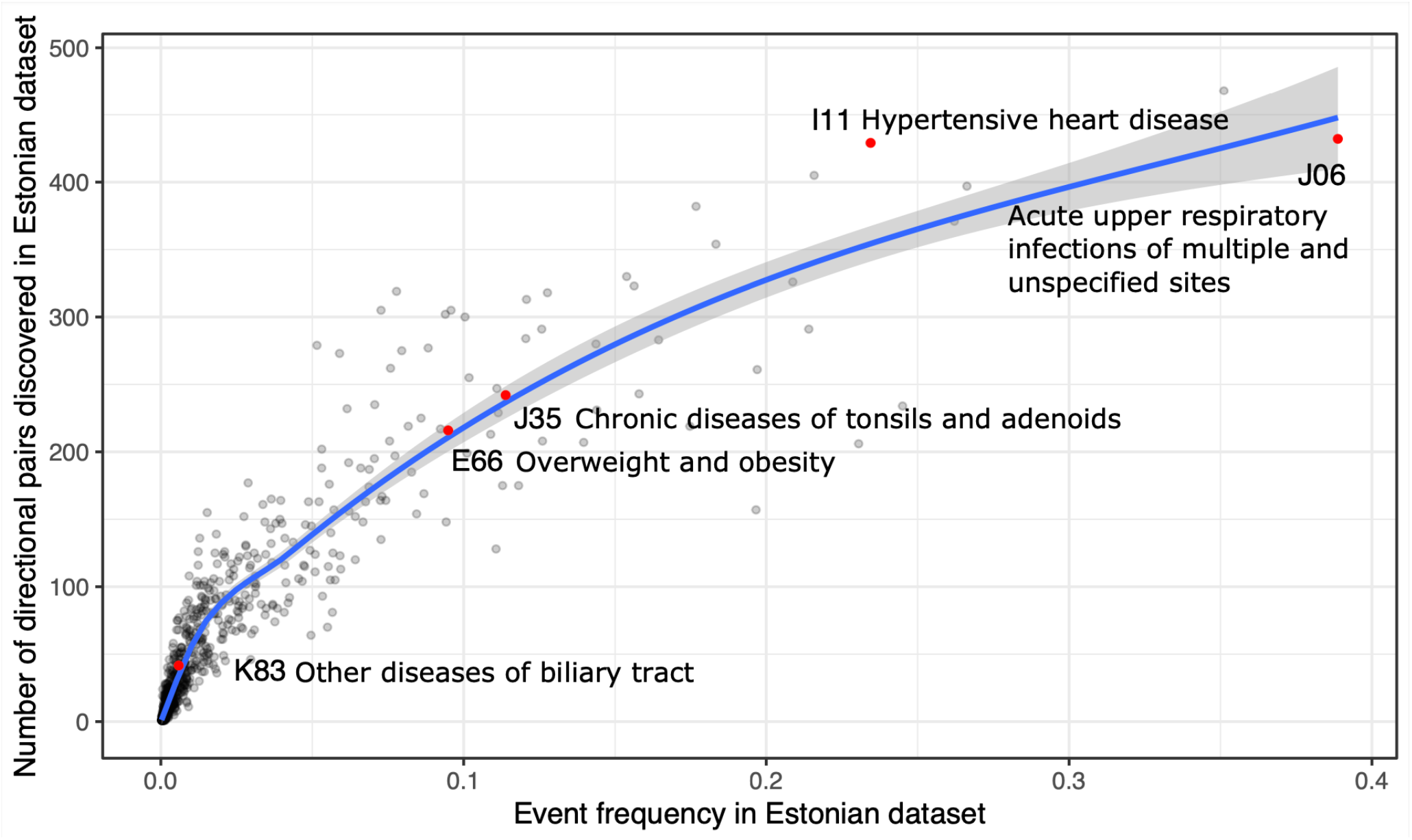
Event frequency is correlated with the number of identified significant event pairs (example on Estonian data). Diagnosis codes mentioned in this paper are highlighted.

All tested and discovered event pairs are given in **Supplementary File**. For privacy reasons, event counts less than 20 are hidden.

### Type 2 Diabetes event trajectories in Estonia and IPCI

To illustrate a cohort-based approach, we ran a discovery study of event trajectories for Type 2 Diabetes (T2D) cohort in the Estonian database (n=11,009 patients) to identify temporal event pairs that occur in at least 1% of the cohort and where the preceding event increases the risk of the second one at least 1.2 times. The discovery run on Estonian data revealed 943 significant event pairs. To validate the findings, we ran a validation study of these pairs on an independent IPCI database from the Netherlands. The validation confirmed 177 of the tested pairs (19%) while 61% of the pairs never occurred in the IPCI database, highlighting the issue of different source codes and/or OMOP CDM mappings used in these databases. In particular, IPCI uses ICPC coding system while Estonian data is based on ICD-10. The latter differentiates T2D with and without complication whereas the IPCI database has a single source code for both conditions (“Type 2 diabetes mellitus”). Therefore, pairs with “Type 2 diabetes mellitus without complication” do not occur there at all. This of course alters the RR values - in IPCI, the sole T2D diagnosis code increases the risk of future metformin 32 times (95%CI 28..37) while in Estonia the risk is much smaller (RR=2.8) as there are other T2D diagnosis codes that precede metformin treatment.

As expected, “Type 2 diabetes mellitus” → “metformin” is the most prevalent trajectory within Estonian data as metformin is the main medication for T2D. Out of longer trajectories, “Essential hypertension” → “Type 2 diabetes mellitus” → “metformin” is the second most prevalent one. This is a somewhat expected result as high blood pressure is a previously shown risk factor for diabetes, especially when the blood pressure is uncontrolled while the order of these conditions also varies in different studies.[14-18] The first most prevalent three-event trajectory is “Type 2 diabetes mellitus” → “metformin” → “metoprolol”, supporting the recent findings of T2D being likely causal to hypertension.[18] T2D damages blood vessels and increases the risk of various cardiovascular diseases,[19] for which metoprolol was the first-line treatment until 2019 in Estonia. In the Netherlands, the most prevalent trajectory is “Cystitis” → “Urinary tract infectious disease”, supporting the previous findings that infectious diseases, including cystitis, are more prevalent among T2D patients when compared to others.[20]

All confirmed event pairs of this study, as well as the trajectories with their counts in the Estonian dataset, are given in **Supplementary File**. These pairs are also available as a built-in preset in the Trajectories package so that everyone can validate those on their own database with only a few clicks. The twenty most frequent pairs are shown in **Figure 2**.

## DISCUSSION

In this paper, we have introduced a framework and implemented an open-source software for detecting event trajectories in OMOP-formatted health data. We evaluated the framework by replicating a Danish study to identify all significant event pairs in Estonia and also conducted a Type 2 Diabetes study on two different datasets.

It can be seen from the results that differences in the frequencies of used event concepts (codes) play an important role in trajectory analysis. There is a strong correlation between the frequency of the event and the number of significant temporal event pairs containing that event (correlation coefficient is 0.88 in Estonia and approximately 0.99 in Denmark^2^). Therefore, when the baseline frequencies vary in different datasets or populations, we will get different sets of significant event pairs. The reasons behind these variations in frequencies can be attributed to several factors. First, the true prevalence of the diseases can be different in different populations or data sets. Second, as we saw above, different concepts can be used in different cultures or healthcare environments to record the same underlying condition. Third, differences in source coding systems and their granularity lead to different mappings and concepts used in OMOP CDM, making them challenging to compare across several databases.[21] If concepts were automatically generalized at a higher level, we might be able to replicate findings more effectively. This would not only resolve the problem of using different concepts but also the issue of low statistical power as the numbers in the case of generalized concepts would be higher. However, as OMOP CDM uses SNOMED Clinical Terms ontology as the underlying vocabulary, moving upwards towards the root of the ontology is a challenging task due to the multiple parent concept principle. Finally, the time span of the dataset also has its implications - the longer the observation period, the more conditions occur (such as chronic diseases or deaths, for example), increasing the frequencies of the events and leading to more event pairs with these events as a result. On the other hand, the longer the observation periods grow, the more age-dependent temporal relationships we can start observing in the data.

Another aspect that needs to be kept in mind is that event trajectories happening in the data and picked up by the software package may not be causative. For instance, confounding effects can cause spurious associations, and it is not easy to distinguish them from the causative event trajectories. The proposed framework does not yet contain any causality checks and the results characterize only the associations in the dataset. However, the findings could be used as hypotheses for further causality studies.

One of the weaknesses of the package is that in its current approach, it is limited to discrete or binary events only. Future extensions of the framework can also include numerical values such as laboratory measurements as events.

There are a number of strengths to our approach as well. While following the principles of previously published studies, it is to our best knowledge the first open-source analysis package for investigating clinical event trajectories. Anyone can now use, examine, validate and modify it as the source code is publicly available. It can be automatically run on any database in OMOP format, not only to characterize the data via trajectories but also to validate event pairs from other studies. The package is not limited to disease codes only as it considers other health-related events such as drug exposures, observations, and procedures, also. The whole analysis process is implemented in a single software package, making each step transparent, and as a whole, stands as a basis for reproducible science. We believe this package can greatly boost scientific studies on the analysis of temporal health events globally and will open new avenues for extending it with additional features in the future.

In the **Supplementary File**, we publish event pairs and disease trajectories from the Estonian population. These can be used as comparison data for any population-level trajectory study in the future.

Finally, we aimed to compare our package results to the output of another disease trajectory tool, recently published by Giannoula et al.[4] This tool detects temporal event pairs and then clusters the trajectories using a dynamic time warping algorithm. To perform the clustering, ICD-9 codes are used. As our dataset does not use ICD-9 codes in OMOP data, we tried to compare only the event pairs detected from both tools. We found the guidelines for using the tool insufficient as we were not able to run the scripts without altering them. Part of the analysis was also missing, such as the code for calculating RR. Attempts to contact the authors have been unsuccessful and therefore, we were unable to compare the performance of these tools. We think this clearly illustrates why open-source pipelines are important.

## CONCLUSION

The proposed framework allows for the identification of significant clinical event progression patterns in health data standardized to the OMOP CDM. We have implemented all of this as an easy-to-use R package Trajectories that enables users to extract and visualize temporal event trajectories from OMOP-formatted observational health data and compare the results across databases.

## Data Availability

All data produced in the present work are contained in the manuscript and supplementary material

## ACKNOWLEDGEMENTS

This work was supported by the Estonian Research Council grants (PRG1095, RITA1/02-96-11); the European Union through the European Regional Development Fund grant EU48684; and the European Social Fund via IT Academy programme. The European Health Data & Evidence Network has received funding from the Innovative Medicines Initiative 2 Joint Undertaking (JU) under grant agreement No 806968. The JU receives support from the European Union’s Horizon 2020 research and innovation programme and EFPIA. We would like to thank Marianna Laht for her great support when providing medical know-how for explaining the results.

## SUPPLEMENTARY DATA

Supplementary File (zip) contains:

- Supplementary Description – Description of Data tables (Supplementary Files 2-4)
- Supplementary Files 2-4 – Data tables

## Footnotes

1 https://github.com/EHDEN/Trajectories

2 Based on 10 random pairs as the exact counts are not published

## REFERENCES

1 Jensen AB, Moseley PL, Oprea TI, et al. Temporal disease trajectories condensed from population-wide registry data covering 6.2 million patients. Nature Communications 2014;5(1):4022.

2 Hu JX, Helleberg M, Jensen AB, et al. A Large-Cohort, Longitudinal Study Determines Precancer Disease Routes across Different Cancer Types. Cancer Res 2019;79(4):864–72.

3 Siggaard T, Reguant R, Jørgensen IF, et al. Disease trajectory browser for exploring temporal, population-wide disease progression patterns in 7.2 million Danish patients. Nature Communications 2020;11(1):4952.

4 Giannoula A, Centeno E, Mayer MA, et al. A system-level analysis of patient disease trajectories based on clinical, phenotypic and molecular similarities. Bioinformatics 2021;37(10):1435–43.

5 Wongsuphasawat K, Guerra Gómez JA, Plaisant C, et al. LifeFlow: visualizing an overview of event sequences. Proceedings of the SIGCHI conference on human factors in computing systems 2011:1747–1756.

6 Monroe M, Lan R, Lee H, et al. Temporal event sequence simplification. IEEE transactions on visualization and computer graphics 2013;19(12):2227–36.

7 Observational Health Data Sciences and Informatics. The Book of OHDSI. https://ohdsi.github.io/TheBookOfOhdsi/ Access date: Nov 2021

8 Sachson C, Ryan P, Kostka K, et al. Our journey. Where The OHDSI Community Has Been And Where We Are Going. https://www.ohdsi.org/wp-content/uploads/2021/09/OHDSI-Book2021-Final.pdf Access date: Nov 2021

9 Chandran U, Reps J, Stang PE, et al. Inferring disease severity in rheumatoid arthritis using predictive modeling in administrative claims databases. PLOS ONE 2019;14(12):e0226255.

10 Reps JM, Schuemie MJ, Suchard MA, et al. Design and implementation of a standardized framework to generate and evaluate patient-level prediction models using observational healthcare data. Journal of the American Medical Informatics Association 2018;25(8):969–75.

11 Li X, Ostropolets A, Makadia R, et al. Characterising the background incidence rates of adverse events of special interest for covid-19 vaccines in eight countries: multinational network cohort study. bmj 2021;373.

12 Stuart EA, King G, Imai K, et al. MatchIt: nonparametric preprocessing for parametric causal inference. Journal of statistical software 2011.

13 Hansen BB, Klopfer SO. Optimal full matching and related designs via network flows. Journal of computational and Graphical Statistics 2006;15(3):609–27.

14 Gress TW, Nieto FJ, Shahar E, et al. Hypertension and antihypertensive therapy as risk factors for type 2 diabetes mellitus. New England Journal of Medicine. 2000;342(13):905–12.

15 Izzo R, De Simone G, Chinali M, et al. Insufficient control of blood pressure and incident diabetes. Diabetes Care 2009;32(5):845–50.

16 Kim MJ, Lim NK, Choi SJ, et al. Hypertension is an independent risk factor for type 2 diabetes: the Korean genome and epidemiology study. Hypertension Research 2015;38(11):783–9.

17 Horr S, Nissen S. Managing hypertension in type 2 diabetes mellitus. Best practice & research Clinical endocrinology & metabolism 2016;30(3):445–54.

18 Sun D, Zhou T, Heianza Y, et al. Type 2 diabetes and hypertension: a study on bidirectional causality. Circulation research 2019;124(6):930–7.

19 Zhang R, Mamza J, Morris T, et al. Lifetime risk of cardiovascular-renal disease in type 2 diabetes: a population-based study in 473399 individuals. European Heart Journal 2020;41(Supplement_2):ehaa946–3313.

20 Shah BR, Hux JE. Quantifying the risk of infectious diseases for people with diabetes. Diabetes care 2003;26(2):510–3.

21 Ostropolets A, Reich C, Ryan P, et al. Characterizing database granularity using SNOMED-CT hierarchy. AMIA Annu Symp Proc. 2021;2020:983–992.

