## Supplementary material for "Trajectories: a framework for detecting temporal clinical event sequences from health data standardized to the OMOP Common Data Model": SupplementaryDescription.docx

### Part 1. Replicating Danish study

In this supplementary document, we assess the disease trajectory detection capability of the Trajectories package by comparing its output on Estonian dataset to the findings described in the paper by Siggaard et al. (<https://www.nature.com/articles/s41467-020-18682-4>). Siggaard et al. uses data from over 7.2 million patients spanning 24 years in Denmark. Unfortunately, we do not have access to this underlying data but we can use the discharge summaries, claims, and prescription data from 10% of a random sample from Estonia, covering 147 thousand patients spanning 8 years (2012-2019).

We run the Trajectories package on Estonian data and compare the results to findings from Denmark. We do this by using two “modes”:

- First, we validate the findings from the Danish dataset on Estonian data
- Second, we detect significant event pairs from the Estonian data and compare the results to the Danish results.

#### Danish data

The data is taken from Supplementary Data 1 of the article by Siggaard et al. (<https://www.nature.com/articles/s41467-020-18682-4>). It contains all associated event pairs in the Danish population that occur at least 20 times. Events are denoted by 3-character ICD-10 code. Note that the data contains an error by using code ‘D99’ instead of ‘Y99’ as far as we understood (the inquiries to the corresponding author of the paper yielded no response). We fixed this error as a first step in our analysis.

There were 77,294 event pairs in the Danish data (where events are associated).

40,711 pairs had significant direction and all these were selected for the validation.

#### Estonian data

The Estonian data covers discharge summaries, insurance claims, and digital prescriptions of a 10% random sample from the Estonian population. Both Estonian and Danish data use ICD-10 diagnosis codes. Insurance claims also contain data about deaths that we use in this analysis, but it is not a comprehensive source for mortality rates. By count, it is estimated that this covers approximately 2/3 of all deaths. All diagnosis codes on the same claim record have the same date which means that events from the same claim are not ordered, even if they actually happened in a specific order (e.g., during in-patient visit).

Similarly to Siggaard et al., we generalized all ICD-10 codes to three-character categories and used a special code ‘Y99’ to denote deaths in the Estonian data. We excluded ICD-10 chapters XVI (P), XVIII (R), XIX (S,T), XX (V,W,X,Y), and XXI (Z) from the analysis as described in the paper by Siggaard et al. Also, we slightly edited the package source code for this specific comparison to be able to handle character-based event codes instead of integer values that are used in OMOP CDM.

For both of the aims described above, the Trajectories package was run by allowing a maximum of 5 years of separation between any two events, and significant directional event pairs were detected.

#### Validating Danish directional event pairs

In the Danish data, there were 40,711 event pairs with significant direction. In this section, we tested out how many of them replicate in Estonian data. The results were as follows:

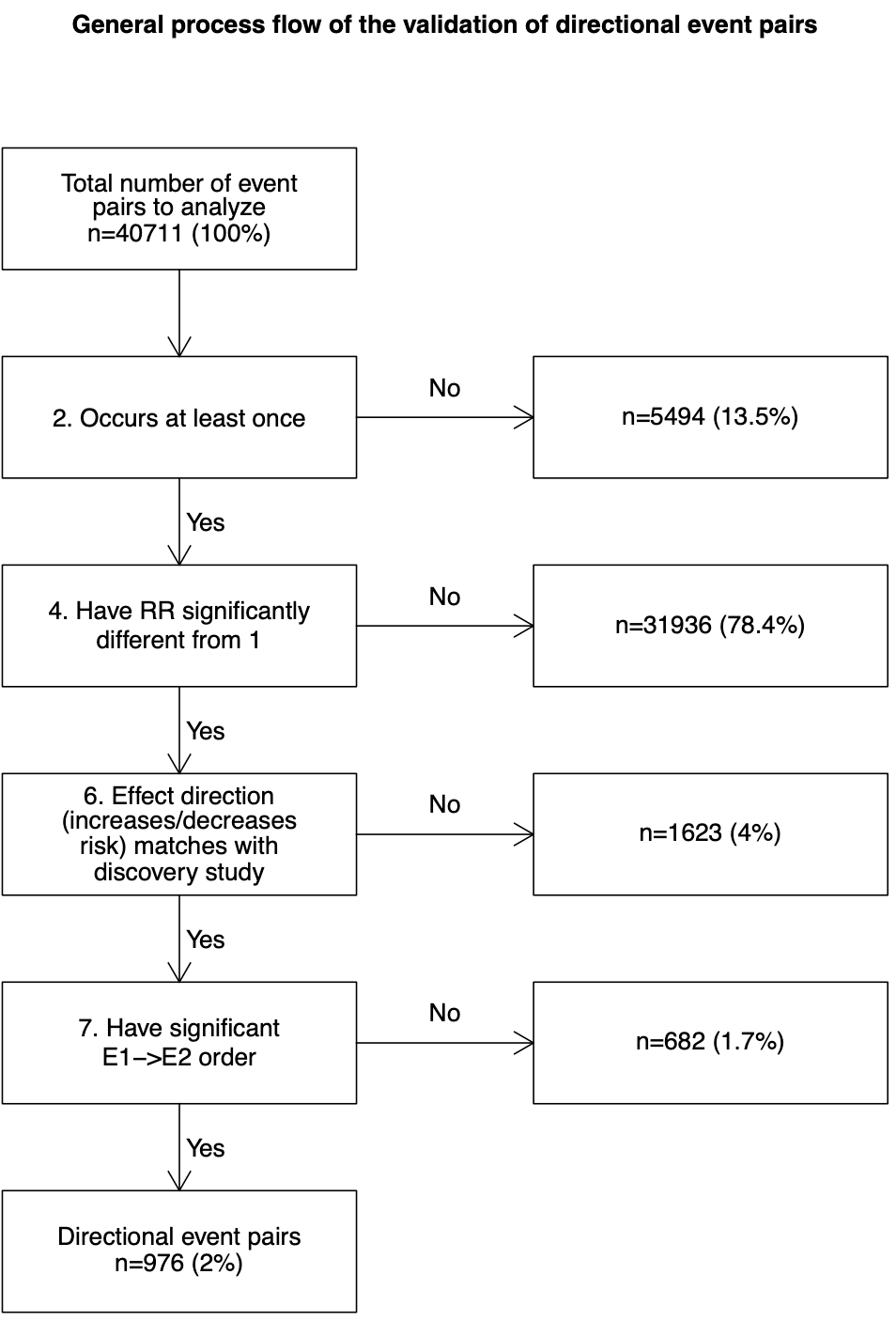

#### Discovering directional event pairs in Estonian data

We used the same minimum patient count >=20 threshold as in Siggaard et al. for the analysis in the Estonian data.

There are 130,137 event pairs in the Estonian data that were tested in total.

Among all tested pairs, events in 63,619 pairs were found to have significant effect (RR) and in 22,618 pairs directionally associated.

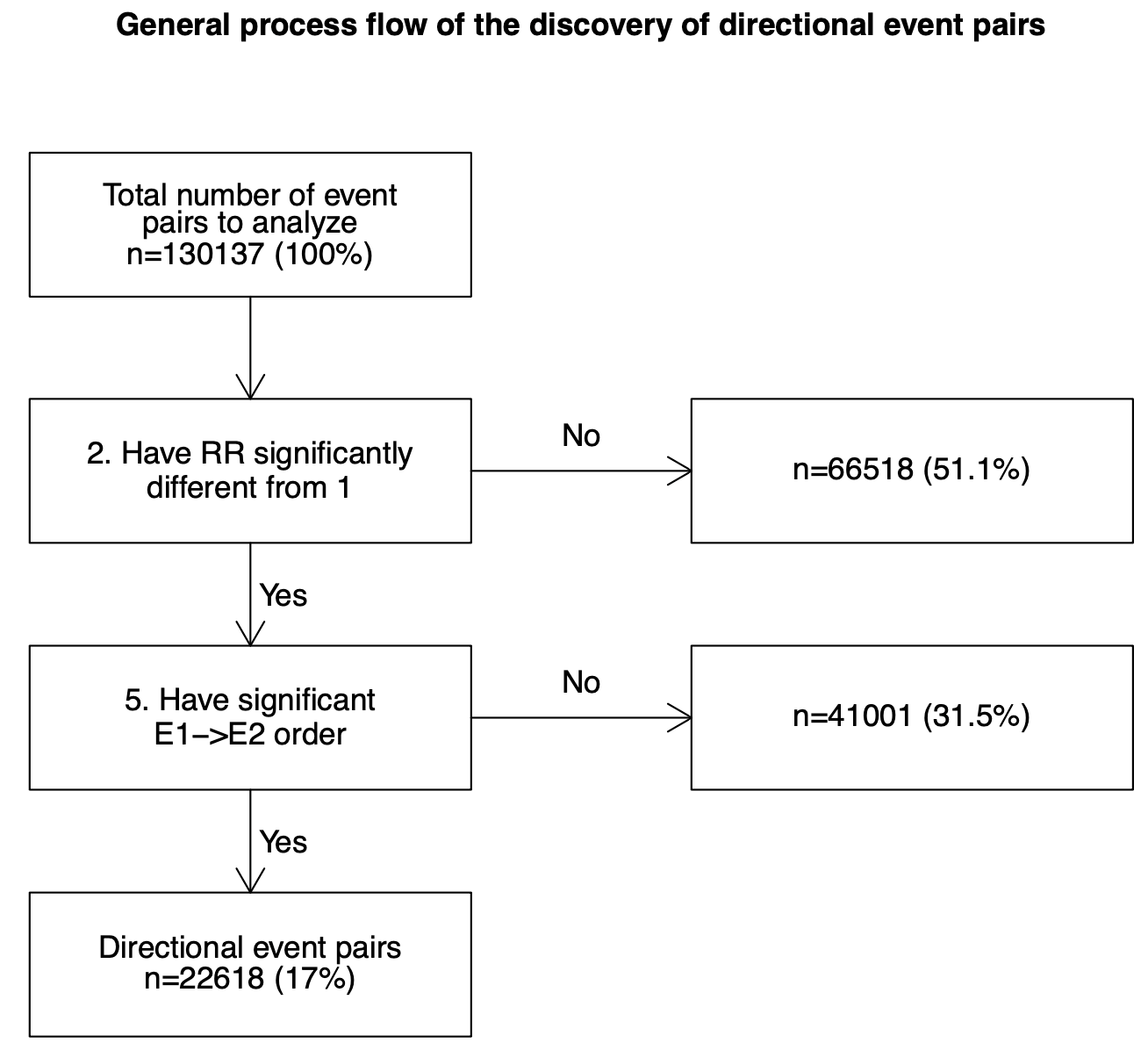

Out of 22,618 directional event pairs detected from Estonian data, 4937 were also published as directional in the Danish population. In 2290 pairs the effect has the same direction.

In Estonian data, event count and the number of directional event pairs are highly correlated (correlation coefficient=0.88)

**The results from both runs are combined in Supplementary File 1.**

### File Name: Supplementary File 1

Description: The Supplementary File 2 contains all ICD-10 diagnosis pairs that were reported directional either in Siggaard et al. 2020 or were discovered directional in Estonian dataset.

Columns:

- EVENT1 - the first diagnosis of the diagnosis pair, encoded in International Statistical Classification of Diseases and Related Health Problems version 10 (ICD-10).
- EVENT2 - the second diagnosis of the event pair
- ASSOCIATED_IN_DENMARK - indicates whether the diagnoses of the pair were reported as associated in Siggaard et al.
- DIRECTIONAL_IN_DENMARK - indicates whether the diagnoses of the pair had a significant temporal order in Siggaard et al.
- RR_IN_DENMARK - Relative risk of the EVENT2 in Siggaard et al.
- PREVALENCE_IN_DENMARK - prevalence of the pair among the total population in Siggaard et al.
- DIRECTIONAL_IN_ESTONIA - indicates whether the diagnoses of the pair had a significant temporal order in Estonia and at the same time the first event significantly alters the risk of the second event
- RR_IN_ESTONIA - Relative risk of the future EVENT2 in Estonia
- PREVALENCE_IN_ESTONIA - prevalence of the pair among the total population of Estonian dataset

### Part 2. Identifying the trajectories of Type 2 Diabetes cohort

In this part, we defined the Type 2 Diabetes study cohort as all patients having any Type 2 Diabetes diagnosis. We then ran the Trajectories package on the Estonian data to detect all trajectories where the preceding event (either condition or drug era) increases the following event at least 1.2 times. Only the pairs that occurred on at least 1% of the cohort were included in the analysis. Also, we set the maximum distance between the events of a pair at 1 year.

We identified 943 event pairs in the Estonian data.

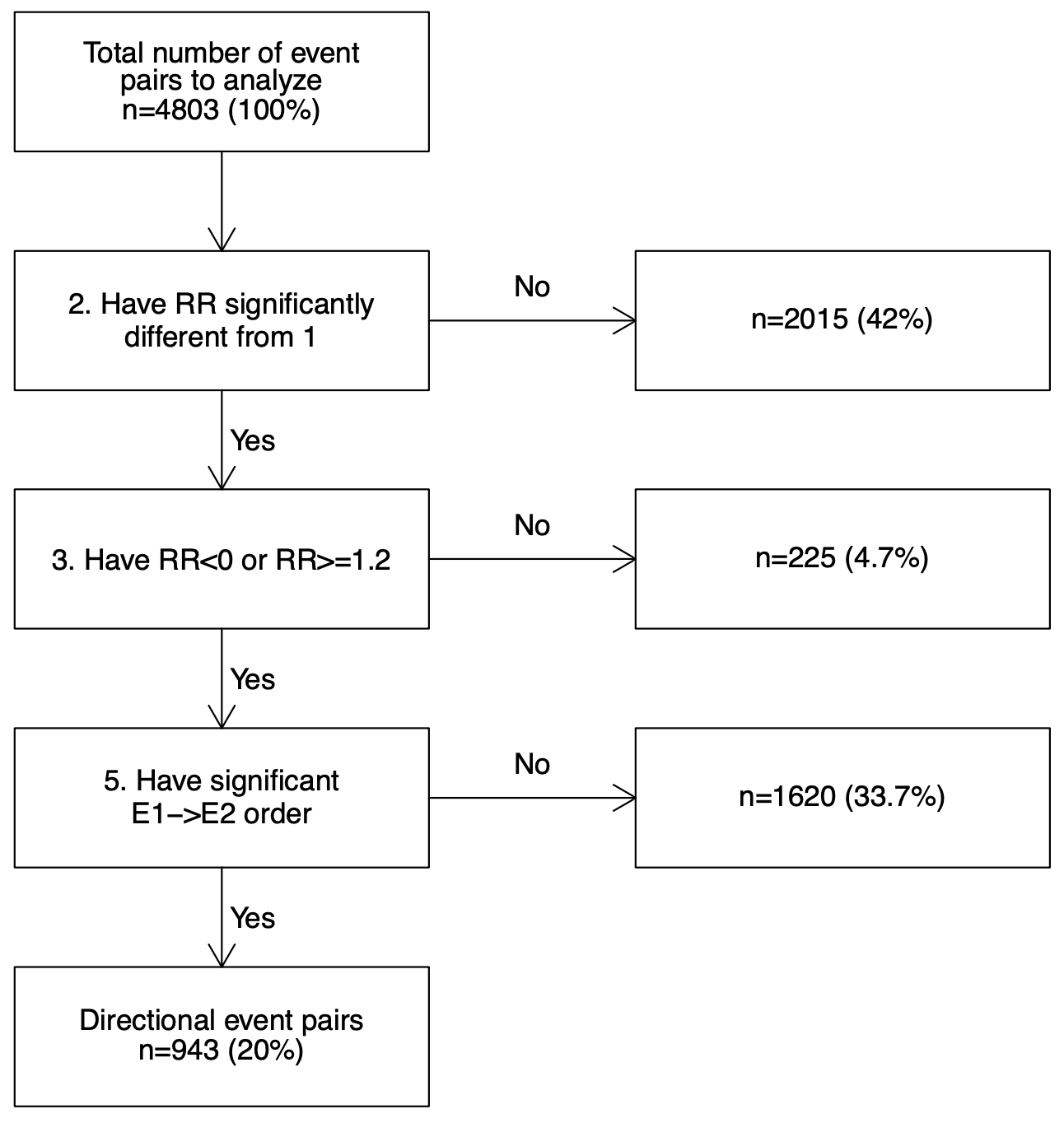

We turned to the IPCI database and let the package validate our previous findings on that data. As a result, for 177 pairs, the effect direction and temporal order of the events were confirmed.

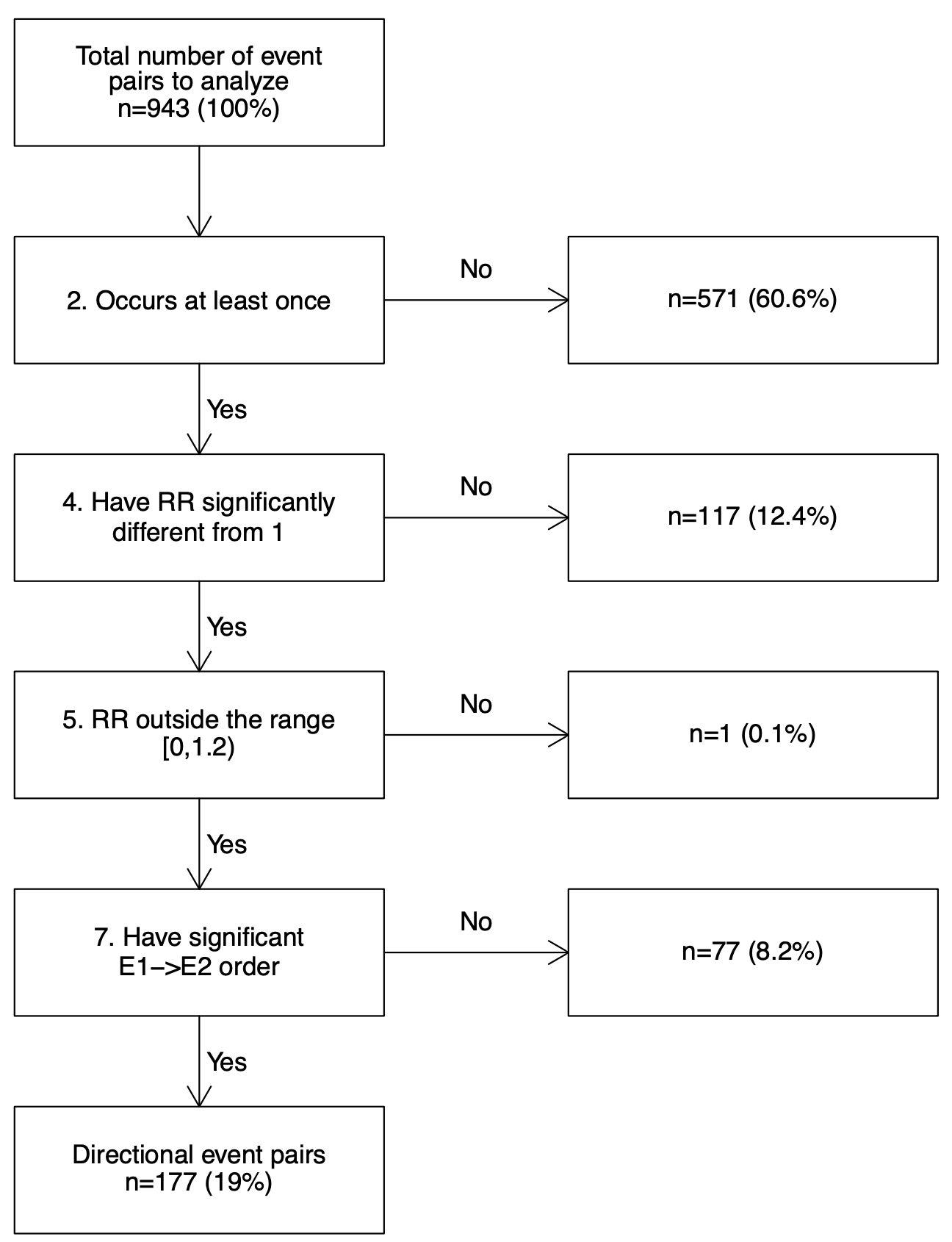

**These 177 pairs are given in Supplementary File 2. The trajectories of these pairs in the Estonian dataset are given in Supplementary File 3.**

### File Name: Supplementary File 2

Description: The Supplementary File 2 contains all event (condition or drug era) pairs where the first event occurred before the second event (statistically significant) and the first event increased the risk of the following second event at least 1.2 times. All these pairs were significant in Estonia and the Netherlands.

Columns:

- E1_CONCEPT_ID - the first diagnosis of the diagnosis pair, encoded by using OMOP CDM standard vocabulary (available at <https://athena.ohdsi.org/search-terms/start>)
- E2_CONCEPT_ID - the second diagnosis of the diagnosis pair, encoded by using OMOP CDM standard vocabulary (available at <https://athena.ohdsi.org/search-terms/start>)
- E1_NAME - the name of the first diagnosis
- E2_NAME - the name of the second diagnosis
- E1_DOMAIN - event domain of the first diagnosis
- E2_DOMAIN - event domain of the second diagnosis
- RR_IN_ESTONIA - Relative risk of the future E2_CONCEPT_ID in Estonia
- RR_IN_IPCI - Relative risk of the future E2_CONCEPT_ID in the Netherlands

### File Name: Supplementary File 3

Description: The Supplementary File 3 contains all trajectories of T2D cohort patients in Estonian dataset where the trajectories are composed of the event pairs given in Supplementary Data 2.

Columns:

- id - id of the trajectory
- trajectory.str - Trajectory as a sequence of concept_id-s, encoded by using OMOP CDM standard vocabulary (available at <https://athena.ohdsi.org/search-terms/start>)
- trajectory.count - The number of patients that had that trajectory in their record
- length - length of the trajectory
- Is_subtrajectory_of - refers to id-s of other trajectories which are longer versions of the current trajectory
- trajectory.str.names - same as “trajectory.str” but the concept_id-s are replaced by concept names
